# Serological responses and six-month trajectories to COVID-19 Comirnaty and Spikevax booster vaccine, September 2021 to January 2022, London, United Kingdom

**DOI:** 10.1101/2022.02.17.22271126

**Authors:** Georgina Ireland, Heather Whitaker, Shamez N Ladhani, Frances Baawuah, Sathyvani Subbarao, Suzanne Elgohari, Alexandra Smith, Michelle O’Brien, Corinne Whillock, Oliver Martin, Paul Moss, Mary E Ramsay, Gayatri Amirthalingam, Kevin E Brown

**Affiliations:** Immunisation and Vaccine Preventable Diseases Division, UK Health Security Agency, London, United Kingdom; Statistics, Modelling and Economics Department, UK Health Security Agency, London, United Kingdom; Brondesbury Medical Centre, Kilburn, London, United Kingdom; Institute of Immunology and Immunotherapy, University of Birmingham, Edgbaston, United Kingdom; Paediatric Infectious Diseases Research Group, St. George’s University of London, London, United Kingdom

**Keywords:** COVID-19, COVID-Vaccine, Antibody, Spike Protein, Immunity, Pfizer, AstraZeneca, Moderna, Comirnaty, Vaxzevria, Spikevax

## Abstract

In contrast to the increasing levels of high avidity S antibody measured by the Roche assay in the first 6 months following natural infection, marked waning is seen post 2 or 3 doses of vaccine. Although the kinetics differ between those with vaccine-induced immunity compared to those infected prior to vaccination (hybrid immunity), waning rates appear to be similar following 2 or 3 doses of vaccine. These data should allow countries to optimise the timing of future doses of vaccine.

---

In September 2021, the UK Joint Committee on Vaccination and Immunisation (JCVI) recommended a third dose (booster) with either a single dose of Comirnaty (BNT162b2 mRNA, BioNTech-Pfizer) or a half dose (50µg) of Spikevax (mRNA-1273, Moderna) for adults aged ≥50 years, individuals aged 16-49 years in clinical risk groups, adult carers and household contacts of immunosuppressed individuals, and frontline health and social care workers, to be offered at least 6 months after their second dose.^1^ Following the emergence of the SARS-CoV-2 Omicron variant in December 2021, the booster vaccine programme was extended to all adults (≥18 years) at least 3 months after second dose due to concerns about waning immunity post-primary vaccination, increased transmissibility of the new variant and the winter pressures on the national healthcare system.^2^

Here we describe antibody kinetics after booster vaccination in adults aged ≥50 years and predict antibody trajectories ≤6 months after booster vaccine. Our findings have important implications for recommendations of further doses of COVID-19 vaccine in boosted adults.

## The CONSENSUS audit

In England, the UK Health Security Agency (UKHSA) initiated an evaluation of vaccine responses in adults aged ≥50 years who received the Comirnaty or Vaxzevria (ChAdOx1-S, AstraZeneca) as part of the national immunisation programme to compare short versus longer interval vaccine schedules and monitor antibody waning over time.^3^ The COVID-19 vacci<u>n</u>e responses after <u>e</u>xtended immunisation <u>s</u>ched<u>u</u>les (CONSENUS) cohort has been described previously.^3,4^ CONSENSUS recruited immunocompetent adults aged ≥50 years in January 2021 in London to provide serial blood samples after their first dose of COVID-19 vaccine. As part of the national COVID-19 vaccine roll out, participants received either (i) two Comirnaty doses <30 days apart (Comirnaty-control), (ii) two Comirnaty doses ≥30 days apart (Comirnaty-extended) or (iii) two Vaxzevria doses ≥30 days apart (Vaxzevria-extended). Additional blood samples were taken before and after booster vaccine (Comirnaty or Spikevax).

Serum samples were tested for nucleoprotein (N) antibodies as a marker of previous SARS-CoV-2 infection (Elecsys Anti-SARS-CoV-2 total antibody assay, Roche Diagnostics, Basel, Switzerland: positive ≥1 COI) and spike (S) protein antibodies, which could be infection- or vaccine-derived (Elecsys Anti-SARS-CoV-2 S total antibody assay, Roche Diagnostics: positive ≥ 0.8 arbitrary units (au)/mL to assess vaccine response). Individuals with ≥0.4 (au)/mL on the N assay were considered to have had prior SARS-CoV-2 infection, this was assessed at enrolment and if participants tested N antibody positive after vaccination, indicating vaccine breakthrough, this and subsequent samples were removed from the analysis.

S antibody geometric mean levels (GMLs) were calculated with 95% confidence intervals (CI). Geometric mean ratios (GMR) of responses between timepoints for each vaccine group were estimated using a mixed regression model on log responses including a random effect for each participant. The GMR by vaccine type at each post-vaccination timepoint was estimated via regression on log Roche S responses and included age-group and sex.

Mixed effects linear regression models were used to model log(S antibody) trajectories over time, including a random effect for each individual. Trajectories were compared to convalescent sera taken up to 400 days after PCR-confirmed infection among unvaccinated 15–98-year-olds. Comparing polynomial models using likelihood ratio tests, for the unvaccinated convalescent data log(days since onset) (from day 21) provided the best fit, while among the vaccinated √(days since 2nd vaccination dose) (from day 14) independently provided the best fit for previously infected and infection-naïve adults. A term for dose was further included with the addition of third Comirnaty dose data, so vaccinated models assume that antibody levels follow a parallel rate of waning after doses 2 and 3. As post-booster data are only available to ∼3 months after dose 3, data on dose 2 responses supplement predictions of longer-term waning after dose 3. Analyses were carried out using STATA v14.2.

### Booster Dose Response

Of the 888 recruited participants, 471 provided serum samples for up to 14 weeks after their booster with Spikevax or Comirnaty (**Table 1**). GMLs increased 2-4 weeks after booster dose for all primary and booster combinations with available data (**Table 2**). In infection-naïve participants, GMLs were 24,901 (95%CI: 10,056-61,663) in Comirnaty-extended/Spikevax, 20,008 (95%CI: 15,463-25,889) in Vaxzevria-extended/Spikevax, 17,703 (95%CI: 13,647-22,965) in Comirnaty-control/Comirnaty, 14,685 (95%CI: 12,611-17,100) in Comirnaty-extended/Comirnaty and 12,015 (95%CI: 13,647-22965) in Vaxzevria-extended/Comirnaty. At 2-4 weeks after the booster GMLs were higher in females but no difference by age was observed. Where pre- and post-booster data were available, GMLs had increased 83.4 fold in Comirnaty-control/Comirnaty, 58.5-fold in Vaxzevria-extended/Comirnaty and 15.3-fold in Comirnaty-extended/Comirnaty vaccinated participants.

**Table 1:** Characteristics of CONSENSUS participants with samples <3 weeks before and up to 14 weeks after Comirnaty or Spikevax booster vaccine. *where N is less than 60 percentages haven’t been calculated*

| Vaccine type<br>(primary/secondary) | N | Dose 1-2 schedule<br>median: days<br>(IQR) | Dose 2-3 schedule<br>median: days<br>(IQR) | Age:<br>median<br>(IQR) | Sex: Female |  | Ethnicity:<br>White |  | Previously<br>infected |  |
| --- | --- | --- | --- | --- | --- | --- | --- | --- | --- | --- |
|  |  |  |  |  | n | % | n | % | n | % |
| Vaxzevria-extended/<br>Comirnaty | 108 | 64 (54 - 76) | 187 (182 - 193.5) | 68 (65 - 71) | 62 | 57.4 | 90 | 83.3 | 17 | - |
| Vaxzevria-extended/ Spikevax | 68 | 77 (70.5 - 77) | 182 (181 - 187.5) | 56 (53 - 59) | 39 | - | 57 | - | 14 | - |
| Comirnaty-extended/<br>Comirnaty | 219 | 76 (73 - 77) | 187 (182 - 193) | 72 (69 - 78) | 124 | 56.6 | 200 | 91.3 | 29 | - |
| Comirnaty-extended/ Spikevax | 10 | 77.5 (76 - 80) | 187 (186 - 203) | 57 (50 - 69) | 9 | - | 10 | - | 0 | - |
| Comirnaty-control/<br>Comirnaty | 66 | 21 (21 - 21) | 262 (261 - 263) | 77.5 (75 - 80) | 33 | - | 61 | 92.4 | 6 | - |

**Table 2:** Geometric mean levels, including 95% confidence intervals, for CONSENSUS participants, including those infected with SARS-CoV-2 prior to vaccination, boosted with Spikevax and Comirnaty vaccines

| Primary | Booster | time<br>sampled<br>after dose 3,<br>weeks | n | geometric mean levels<br>(95% CI) | within-individual<br>geometric mean<br>ratio of response<br>relative to 2-4<br>wks post dose 3<br>(95% CI) | within-individual<br>geometric mean<br>ratio of response<br>relative to <3<br>weeks before dose<br>(95% CI) |
| --- | --- | --- | --- | --- | --- | --- |
| Vaxzevria | Spikevax | 2-4 weeks | 49 | 20008 (15463 - 25889) |  |  |
|  |  | 5-9 weeks | 5 | 20048 (10705 - 37545) |  |  |
|  | Comirnaty | <3 before | 50 | 208 (150 - 289) |  | ref |
|  |  | 2-4 weeks | 63 | 12015 (9651 - 14958) | ref | 58.5 (40.3 - 85) |
|  |  | 5-9 weeks | 15 | 9525 (6354 - 14279) | 0.88 (0.71 - 1.09) |  |
|  |  | 10-14 weeks | 59 | 5209 (4311 - 6295) | 0.51 (0.47 - 0.56) |  |
| Comirnaty,<br>schedule < 30<br>days | Comirnaty | <3 before | 45 | 194 (138 - 272) |  | ref |
|  |  | 2-4 weeks | 47 | 17703 (13647 - 22965) | ref | 83.4 (63.1 - 110.2) |
|  |  | 5-9 weeks | 2 |  |  |  |
|  |  | 10-14 weeks | 49 | 8926 (6383 - 12482) | 0.6 (0.55 - 0.65) |  |
| Comirnaty | Spikevax | 2-4 weeks | 5 | 24901 (10056 - 61663) |  |  |
|  |  | 5-9 weeks | 5 | 17466 (8728 - 34954) |  |  |
|  | Comirnaty | <3 before | 120 | 964 (813 - 1145) |  | ref |
|  |  | 2-4 weeks | 133 | 14685 (12611 - 17100) | ref | 15.3 (13 - 18) |
|  |  | 5-9 weeks | 35 | 11738 (9431 - 14611) | 0.77 (0.66 - 0.91) |  |
|  |  | 10-14 weeks | 149 | 6305 (5572 - 7134) | 0.45 (0.41 - 0.48) |  |
| Infection before vaccination |  |  |  |  |  |  |
| Vaxzevria | Spikevax | 2-4 weeks | 13 | 40135 (27773 - 57998) |  |  |
|  | Comirnaty | <3 before | 12 | 3804 (1253 - 11547) |  | ref |
|  |  | 2-4 weeks | 7 | 49463 (33658 - 72689) | ref | 13.5 (8.3 - 21.9) |
|  |  | 5-9 weeks | 5 | 25056 (10498 - 59802) | 0.77 (0.58 - 1.03) |  |
|  |  | 10-14 weeks | 13 | 12800 (8517 - 19238) | 0.52 (0.45 - 0.61) |  |
| Comirnaty,<br>schedule < 30<br>days | Comirnaty | <3 before | 6 | 8806 (1309 - 59254) |  | ref |
|  |  | 2-4 weeks | 4 | 33130 (6383 - 171950) | ref | 5 (0.9 - 28.1) |
|  |  | 5-9 weeks | 0 |  |  |  |
|  |  | 10-14 weeks | 5 | 17823 (4926 - 64495) | 0.82 (0.78 - 0.85) |  |
| Comirnaty | Spikevax | 2-4 weeks | 2 |  |  |  |
|  | Comirnaty | <3 before | 15 | 5872 (3144 - 10968) |  | ref |
|  |  | 2-4 weeks | 18 | 21278 (14190 - 31908) | ref | 5.3 (3.5 - 8) |
|  |  | 5-9 weeks | 2 |  |  |  |
|  |  | 10-14 weeks | 20 | 15537 (10791 - 22370) | 0.62 (0.53 - 0.72) |  |

GMLs not calculated where 2 or less people within grouping

Additionally, 66 participants had infection with Wuhan or Alpha before primary vaccination and their GMLs were higher than infection-naïve participants, but post-booster GMRs were greater among the latter. In those with past infection GMLs were highest after Vaxzevria-extended/Comirnaty (49,463, 95%CI:33,658-72,689), followed by Vaxzevria-extended/Spikevax (40,135, 95%CI:27,773-57,998), Comirnaty-control/Comirnaty (33,130, 95%CI: 6,383-17,1950) and Comirnaty-extended/Comirnaty (21,278, 95%CI:14,190-31,908), with GMLs increasing 13.5-fold in Vaxzevria-extended/Comirnaty and 5-fold each for Comirnaty-control/Comirnaty and Comirnaty-extended/Comirnaty participants.

### Post-booster trajectories

For all participants, GMLs peaked 2-4 weeks post-booster and started waning from 5-9 weeks. At 10-14 weeks, GMLs had reduced 65% in Comirnaty-extended/Comirnaty, 49% in Vaxzevria-extended/Comirnaty and 40% in Comirnaty-control/Comirnaty in infection-naïve participants and by 48% in Vaxzevria-extended/Comirnaty, 38% in Comirnaty-extended/Comirnaty and 18% in Comirnaty-control/Comirnaty in previously infected participants (**Table 2, Figure 1&2**). The post-second and booster dose S-antibody trajectories in infection-naïve and previously-infected participants following Comirnaty-extended primary plus booster differed, with the relative rate of post-second dose decline lower in those who were previously-infected (day 14:182 ratio: 9.5 and 6.5 respectively). Within the infection-naïve and previously-infected cohorts, however, the 3-month rate of decline was similar after second and booster doses (**Figure 2b-d**). Assuming the same rate of decline as after the second dose, models estimate that at 6 months post-booster vaccine, GMLs would reduce to 2,519 (95%CI: 2,230-2,846) in infection-naïve and 5,945 (95%CI: 4,387-8,057) in previously infected participants.

**Figure 1:**
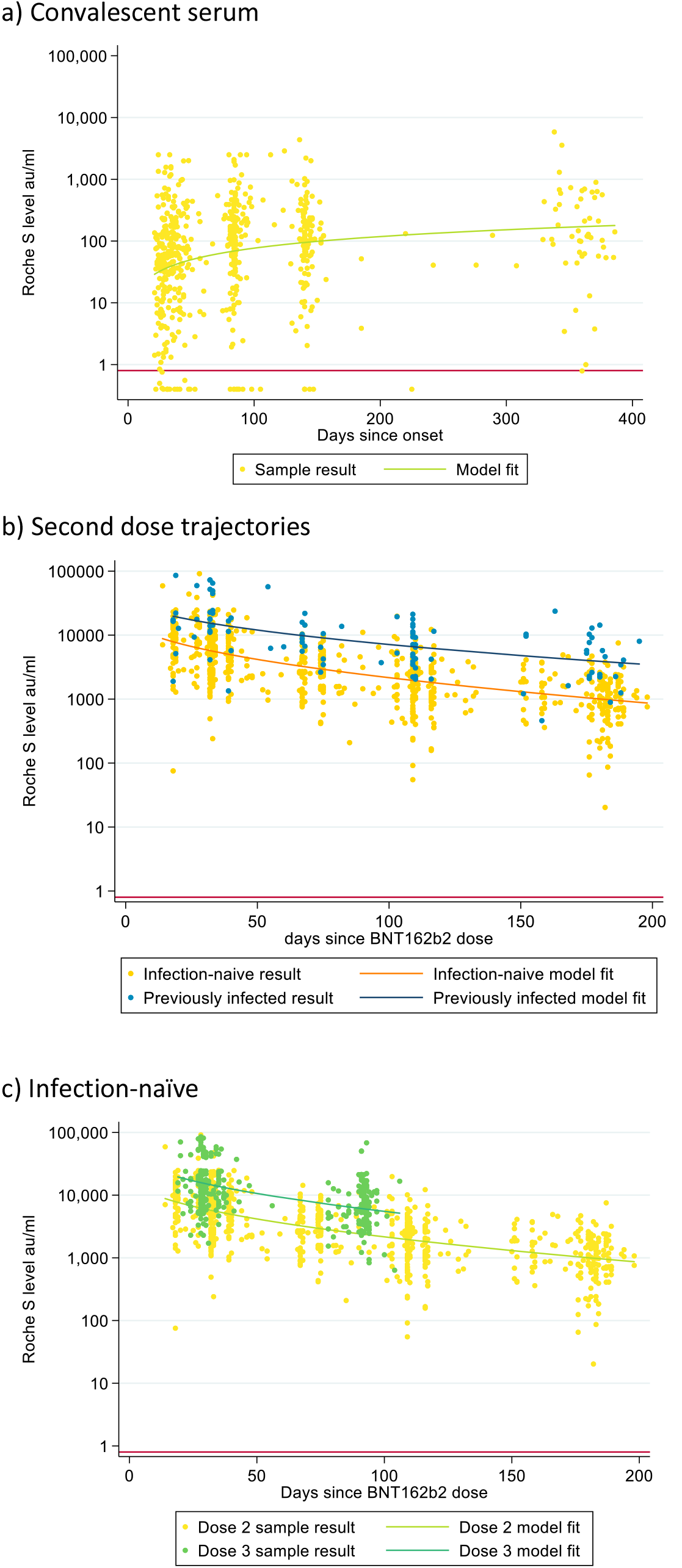
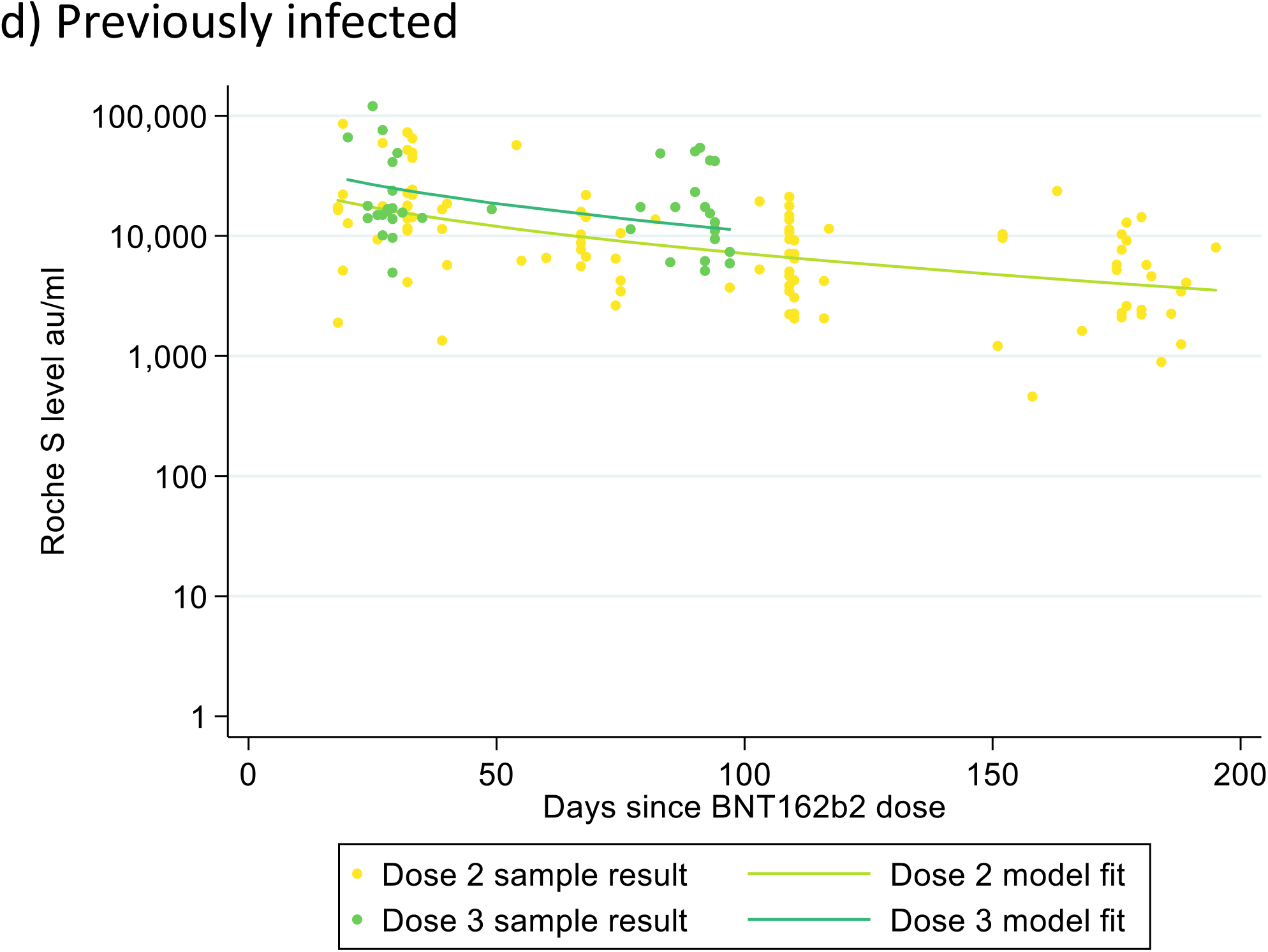
S antibody trajectory after infection in convalescent serum samples (a), after the second vaccine dose with Comirnaty in CONSENSUS participants by prior infection status (b), and after the second and third vaccine dose with no past SARS-CoV-2 infection before vaccination (c) and with a past infection before vaccination (d).

Whilst post-vaccine antibody levels fell after all vaccine combinations, convalescent sera from clinically mild-to-moderate PCR-confirmed cases (due to Wuhan infecting strain) show that S-antibody GMTs increase for up to 400 days post-infection (**Figure 2a**). However, GMTs in convalescent sera are significantly lower than post-vaccine, being 172 (95%CI: 132-225) at 365 days post-infection.

### Implications of findings

Our results demonstrate a rapid serological response post-booster for all primary immunisation vaccines and schedules, to higher levels than observed after dose 2 of the primary schedule, but waning was observed after 5 weeks. By comparison, there was very little S-antibody waning in naturally-infected, unvaccinated adults up to 365 days after infection. Whilst serological assessment does not take cellular or innate immunity into account, S-antibody responses mirror vaccine efficacy analysis against SARS-CoV-2 infection, hospitalisations and death, even with the emergence of Omicron variant.^5,6^ Additionally, post-booster GMTs in our cohort were similar to those observed in COV-BOOST, a study analysing the vaccine responses to different vaccine combinations.^7^

Our data also highlight differences in people with hybrid-immunity (infection and vaccine) who had higher GMTs at all time-points and less S-antibody waning post-booster. Further work is required to understand whether trajectories differ in people with post-vaccine breakthrough infections, and are dependant on the variant of infection.

The rapid waning of post-booster antibodies along the same trajectory as two primary immunisation doses indicates that a fourth dose may be required when predictions of future waves coincide with declining protection, particularly in vulnerable groups. However, we do not know the level above which infections or severe disease are prevented. Countries need to decide whether to offer this dose to all adults, as recently announced in Israel, or for higher-risk groups only, such as older adults and those with comorbidities.^8^ Our trajectory and six-month GML estimates may have utility for policy-makers deciding on the optimal interval for recommending the fourth dose of vaccine, depending on the aim of the programme.

## Data Availability

Applications for relevant anonymised data should be submitted to the UK Health Security Agency Office for Data Release.

## Acknowledgements

We would like to thank Dorothy Blundell, Dr Caroline Sayer and the team at Haverstock Healthcare GP Federation and the whole CONSESUS and Convalescent sera surveillance team at UK Health Security Agency (UKHSA) including those at Colindale, Manchester and Porton Down who assisted with the laboratory testing.

